# Violence and Non-fatal Injuries among Vietnamese In-school Adolescents: National Prevalence Estimates and Associated Factors

**DOI:** 10.1101/2021.04.12.21255309

**Authors:** Phuong Anh Le, Van Minh Hoang, Thi Tuyet Hanh Tran, Quynh Long Khuong, Momoe Takeuchi, Tuan Lam Nguyen, Thi Quynh Nga Pham, Van Tuan Le, Quoc Bao Tran, Kidong Park

## Abstract

**Background:** School violence and injury are major public health problems worldwide; however, current information on these issues in Vietnam is lacking. We aim to investigate the prevalence of violence and non-fatal injury and associated factors among Vietnamese adolescents aged 13-17 years old.

**Methods:** We used data from the 2019 Vietnam Global School-based Student Health Survey (GSHS), which is a nationally representative survey developed by the World Health Organization to monitor behavioral risk factors among school-aged students. The Vietnam GSHS 2019 was conducted in 20 provinces and cities, with a sample size of 7,690 male and female students aged 13-17 years old.

**Results:** We found the prevalence of violence and non-fatal injury was 14.5% and 21.4%, respectively. Common risk factors for both violence and non-fatal injuries included cigarette smoking, alcohol use, mental health problems, and living with neither parent; violence was also a risk factor for non-fatal injuries. Student older age was associated with lower odds of school violence. Parents played an important role in preventing violence among female students and non-fatal injuries in both genders.

**Conclusions:** Future policies should consider individual factors as well as adolescent-parent bonding, to mitigate the burden of violence and injury among in-school adolescents in Vietnam.

## Introduction

School violence occurs when a student is physically attacked or involved in a physical fight at school, on the way to/from school, or at school-related events. Every year, 200,000 homicides occur among young people aged 10-29 years old worldwide.^1^ Furthermore, violence has serious, life-long impacts on physical health, psychological health, and the social functioning of young people.^1^ In a study of student victimization across 37 nations, an average of 27.8% of students reported that they had been victims of school violence at least once in the month preceding the survey. Countries with the highest rates of victims of school violence are Hungary, Romania, and the Philippines.^2^ The prevalence of violence among 68 low-to-middle-income countries (LMICs) is 35.6%.^3^ Indeed, school violence is prevalent in many countries around the world, which makes it a global public health concern.

Serious non-fatal injuries result from various causes, including violence. The classification of serious injury is when an individual is forced to miss at least one full day of normal activities (such as school, sports, or work), or requires treatment by a doctor or a nurse. These injuries lead to more than 5 million deaths annually and are a major public health problem.^4^ Moreover, injury is among the leading causes of death among adolescents worldwide.^5^ A research study that looked at 11-, 13-, and 15-year-old adolescents in 11 industrialized countries, reported that 41.3% of all children had been injured and needed medical treatment in the preceding 12 months.^6^ Among 68 LMICs, serious injury affected 47.8% of male students and 37.5% of female students.^3^ The Global School-based Student Health Survey (GSHS) in four Southeast Asian Countries reported an average of 42.2% of adolescents having suffered one or more serious injuries within the preceding 12 months.^7^

The current literature reports that, among students, school violence is associated with suicide ^8,9^; injury is associated with poor mental health^10-13^ and violence;^9,11,13-15^ and both violence and injury are associated with substance use^7-9,11,16^ and truancy.^9-11,17^ Insufficient parental supervision^9^ and lack of parental bonding^13^ are risk factors for violence and injury, while parental respect and parental support are protective factors.^8,17^

Vietnam is a low-to-middle-income country in the Southeast Asia region, with a population of 95.5 million.^18^ According to GSHS 2013, 34.3% of male middle-school students, and 25.1% of female middle-school students in Vietnam, experienced one or more serious injuries.^19^ Although there are some published papers on violence and injury in industrialized countries and on the overall status in LMICs, scientific evidence on violence or injury and associated factors in Vietnam is very scarced, since school violence is a sensitive topic in Vietnamese culture. In Vietnam, schools with a high prevalence of violence are likely to suffer from negative media coverage and gain a bad reputation; therefore, school officials may have little incentive to facilitate research on school violence. GSHS 2019 was supported by both the Ministry of Health and the Ministry of Education and Training in Vietnam, which facilitated data collection in schools across the whole country and allowed preparation of a nationally representative report of both violence and injury among Vietnamese school students.

In this study, we used data from GSHS 2019 to report on the status of these two topics in Vietnam and to provide the most up-to-date information on the prevalence of violence and non-fatal injury in Vietnam in 2019 and their associated factors.

## Methods

### Data source

This study used data resulting from the Vietnam GSHS 2019, which is a nationally representative survey of students aged 13-17. Hanoi University of Public Health conducted this survey in collaboration with the General Department of Preventive Medicine, the Ministry of Health, and the Ministry of Education and Training, Vietnam. The World Health Organization (WHO) and Center for Disease Control (CDC) provided financial and technical assistance. GSHS 2019 consists of core questionnaire modules with expanded and country-specific questions that cover key areas of behavioral risk factors and protective factors among school-age adolescents.

### Study setting and participants

GSHS 2019 followed the WHO standardized study design, using cross-sectional two-stage cluster sampling to obtain a representative sample of students in senior secondary schools (grades 8-9) and high schools (grades 10-12). The Vietnam GSHS 2019 was conducted in 20 provinces and cities. In the first stage, we selected schools based on the probability proportional to size sampling technique. To make the sample representative for subpopulations, we stratified the sample by place of residence (rural/urban) and school level (senior secondary/high school). We selected a sample size of 1,500 and a response rate of 90% for each stratum, to ensure a margin of error of less than 5%. In the second stage, we applied a simple random sampling technique to choose classes from each selected school. All students in the selected classes were eligible to participate in the study regardless of their actual age. In total, we collected data from 7,796 students in 81 schools and 210 classes. The school and student response rate was 96.4% and 97.0%, respectively.

### Study instruments

We used the WHO standardized GSHS questionnaire to develop the Vietnam-specific instrument. Firstly, we applied forward and backward translation of the English version. Some country-specific questions were then added to form the initial version of the Vietnam GSHS 2019 questionnaire. The instrument was pre-tested in the target population at two schools in Hanoi, Vietnam. After the pre-test, the questionnaire was revised to ensure that students could understand and complete the survey within the allocated time. These students were excluded from the final sample.

### Data collection

Before collecting data at the selected classes, we obtained written informed consent from the students’ parents/guardians with the assistance of the school administrators. In the ensuing days, we surveyed the students who had a completed consent form from their parents/guardians and who were willing to participate in the study. Students answered the self-administered questionnaire on separate, computer scannable answer sheets, during one classroom period.

### Study variables

**Table 1** provides the variable definitions of this study.

**Table 1:** Variable definitions.

| Variables | GSHS 2019 questionnaire | Definition used in this paper |
| --- | --- | --- |
| **Dependent variables** |  |  |
| Violence | - During the past 12 months, how many times were you physically attacked?<br>- During the past 12 months, how many times were you in a physical fight?<br>*1 "0 times" to 8 "12 times"* | Students who were physically attacked or involved in physical fight at least 1 time during the past 12 months<br>*Yes vs. No* |
| Injury | During the past 12 months, how many times were you seriously injured?<br>*1 "0 times" to 8 "12 times"* | Students who were seriously injured at least 1 time during the past 12 months<br>*Yes vs. No* |
| **Independent variable** |  |  |
| Gender |  | *Male vs. Female* |
| Age |  | *13 – 17* |
| Place of residence |  | *Rural vs. Urban* |
| Living with | Do you live with both of your parents? | *Both parents, Mother or father, or Neither* |
| Parents monitoring | - During the past 30 days, how often did your parents or guardians check to see if your homework was done?<br>- During the past 30 days, how often did your parents or guardians really know what you were doing with your free time?<br>*1 "Never" to 5 "Always"* | Students whose parents most of the time/always checked homework or know what their children do in their free time<br>*High vs. Low* |
| Parents respect | - During the past 30 days, how often did your parents or guardians go through your things without your approval?<br>- During the past 30 days, how often did your parents or guardians not respect you as a person (for example, not let you talk or favor someone else more than you)?<br>*1 "Never" to 5 "Always"* | Students whose parents most of the time/always respect their children or their personal space<br>*High vs. Low* |
| Parents support | - During the past 30 days, how often did your parents or guardians understand your problems and worries?<br>- During the past 30 days, how often did your parents or guardians give you advice and guidance?<br>*1 "Never" to 5 "Always"* | Students whose parents most of the time/always understood problems/worries or gave advice/guidance<br>*High vs. Low* |
| Parents expectation | - During the past 30 days, how often do your parents or guardians expect too much of you (for example, to do better in school or be a better person)?<br>*1 "Never" to 5 "Always"* | Students whose parents most of the time/always expect too much on their children<br>*High vs. Low* |
| Number of close friends | How many close friends do you have? | *0, 1, 2, 3 or more* |
| Alcohol use | How old were you when you had your first drink of alcohol?<br>*1 "I have never had a drink of alcohol", 2 "I have never had a drink of alcohol other than a few sips", 3 "7 years old or younger" to 9 "18 years old or older"* | Students who has ever drunk alcohol (rather than a few sips)<br>*Yes vs. No* |
| Smoke cigarettes | How old were you when you first tried a cigarette?<br>*1 "I have never smoked cigarettes", 2 "7 years old or younger" to 8 "18 years old or older"* | Students who has ever smoked cigarettes<br>*Yes vs. No* |
| Truancy | During the past 30 days, on how many days did you miss classes or school without permission?<br>*1 "0 days" to 5 "10 days or more"* | Students who missed any days without permission during the past 30 days<br>*Yes vs. No* |

| **Variables** | **GSHS 2019 questionnaire** | **Definition used in this paper** |
| --- | --- | --- |
| Loneliness | During the past 12 months, how often have you felt lonely?<br>*1 "Never" to 5 "Always"* | Students who most of the time/always felt lonely during the past 12 months<br>*Yes vs. No* |
| Worried | During the past 12 months, how often have you been so worried about something that you could not eat or did not feel hungry?<br>*1 "Never" to 5 "Always"* | Students who most of the time/always felt worry during the past 12 months<br>*Yes vs. No* |
| Suicide attempt | During the past 12 months, did you ever seriously consider attempting suicide?<br>*1 "Yes" to 2 "No"* | Students who seriously considered attempting suicide<br>*Yes vs. No* |

#### Outcome variables

The two outcome variables of the study are violence and non-fatal injuries. Violence was assessed by asking students if they had experienced any physical attacks or been involved in any physical fight in the preceding 12 months. Non-fatal injuries were assessed by asking students if they had been seriously injured in the previous 12 months.

#### Independent variables

We assessed the potential correlates of violence and non-fatal injuries, including demographic characteristics (gender, age, place of residence, living with parents); parental factors (parental monitoring, parental support, parental respect, parental expectation); number of close friends; risk behaviors (alcohol use and the smoking of cigarettes); mental health problems (suicide attempt, loneliness, worrying); and truancy. The details on the variable definitions are shown in Table 1.

### Data analysis

We used frequency and weighted percentages to describe the categorical variables: weighted percentage and 95% confidence interval (CI) to compare the prevalence of violence and injury across groups. Since the patterns of violence and injury are potentially different between gender groups, we stratified all analyses by gender. In this study, among 7,796 students, 106 self-identified as other-gender. Due to the small sample of the other-gender group, we included only male and female gender (n = 7,690) in the stratified analyses.

To identify factors related to the outcomes of interest, we applied multivariate logistic regression. The variables included in the multivariate models were based on our literature review of potential covariates of violence and injury among school-aged adolescents. We utilized the survey package *“svy”* in Stata version 16 SE (StataCorp, College Station, TX, USA) to account for the cluster sampling design and sampling weights of the GSHS. We reported the odds ratio (OR) and 95% CI for modeling results and used a significance level of 0.05 for all statistical tests.

## Results

### Participant characteristics

**Table 2** describes the characteristics of the study participants. More than half of the students were female (54.0%), attending secondary school (55.5%), and living in rural areas (63.5%). Most of the participants lived with both their mother and father (72.4%). Regarding parental factors, 47.7% of parents were involved in their children’s homework and free time, 55.8% understood and gave advice and guidance, 85.7% had respect, and 54.8% had a high expectation of their children. About two-thirds of the students had three or more close friends, 44.0% drank alcohol, and 8.2% smoked cigarettes. Regarding mental health, 12.3% of the students felt lonely, 6.1% felt worried, and 15.4% had attempted suicide in the preceding 12 months. Around 15.1% of the students had committed truancy in the previous 30 days.

**Table 2:** Participants’ characteristics.

| Characteristics | Total |  | Male |  | Female |  |
| --- | --- | --- | --- | --- | --- | --- |
|  | n | Weighted % | n | Weighted % | n | Weighted % |
| **N** | 7690 |  | 3572 |  | 4118 |  |
| **Age** |  |  |  |  |  |  |
| 13 | 1024 | 16.7 | 441 | 15.7 | 583 | 17.6 |
| 14 | 1523 | 30.4 | 684 | 28.6 | 839 | 31.9 |
| 15 | 1672 | 19.9 | 818 | 21.7 | 854 | 18.3 |
| 16 | 1646 | 15.9 | 753 | 16.1 | 893 | 15.7 |
| 17 | 1825 | 17.1 | 876 | 17.9 | 949 | 16.5 |
| **Education level** |  |  |  |  |  |  |
| Senior secondary school | 2940 | 55.5 | 1371 | 55.6 | 1569 | 55.4 |
| High school | 4750 | 44.5 | 2201 | 44.4 | 2549 | 44.6 |
| **Place of residence** |  |  |  |  |  |  |
| Rural | 3833 | 63.5 | 1755 | 62.9 | 2078 | 64.1 |
| Urban | 3857 | 36.5 | 1817 | 37.1 | 2040 | 35.9 |
| **Living with** |  |  |  |  |  |  |
| Both parents | 5608 | 72.4 | 2613 | 73.1 | 2995 | 71.8 |
| Mother or father | 798 | 10.3 | 357 | 9.8 | 441 | 10.7 |
| Neither | 1280 | 17.3 | 599 | 17.1 | 681 | 17.5 |
| **Parental monitoring** |  |  |  |  |  |  |
| Low | 4252 | 52.3 | 1829 | 48.2 | 2423 | 55.7 |
| High | 3410 | 47.7 | 1726 | 51.8 | 1684 | 44.3 |
| **Parental support** |  |  |  |  |  |  |
| Low | 3586 | 44.2 | 1565 | 41.5 | 2021 | 46.5 |
| High | 4102 | 55.8 | 2006 | 58.5 | 2096 | 53.5 |
| **Parental respect** |  |  |  |  |  |  |
| Low | 1176 | 14.3 | 600 | 15.7 | 576 | 13.2 |
| High | 6451 | 85.7 | 2929 | 84.3 | 3522 | 86.8 |
| **Parental high expectation** |  |  |  |  |  |  |
| Low | 3439 | 45.2 | 1529 | 43.1 | 1910 | 47.1 |
| High | 3914 | 54.8 | 1879 | 56.9 | 2035 | 52.9 |
| **Number of close friends** |  |  |  |  |  |  |
| 0 | 778 | 8.8 | 353 | 8.5 | 425 | 9.0 |
| 1 | 1075 | 13.0 | 397 | 10.4 | 678 | 15.2 |
| 2 | 1172 | 15.6 | 457 | 13.0 | 715 | 17.8 |
| ≥3 | 4569 | 62.7 | 2318 | 68.2 | 2251 | 58.0 |
| **Drink alcohol** |  |  |  |  |  |  |
| No | 3987 | 56.0 | 1730 | 53.6 | 2257 | 58.0 |
| Yes | 3659 | 44.0 | 1818 | 46.4 | 1841 | 42.0 |
| **Smoke cigarettes** |  |  |  |  |  |  |
| No | 6954 | 91.8 | 3032 | 86.7 | 3922 | 96.0 |
| Yes | 713 | 8.2 | 525 | 13.3 | 188 | 4.0 |
| **Loneliness** |  |  |  |  |  |  |
| No | 6589 | 87.7 | 3088 | 88.3 | 3501 | 87.2 |
| Yes | 1098 | 12.3 | 481 | 11.7 | 617 | 12.8 |
| **Worried** |  |  |  |  |  |  |
| No | 7133 | 93.9 | 3367 | 95.2 | 3766 | 92.7 |
| Yes | 548 | 6.1 | 198 | 4.8 | 350 | 7.3 |
| **Suicide attempt** |  |  |  |  |  |  |
| No | 6333 | 84.6 | 3082 | 88.1 | 3251 | 81.6 |
| Yes | 1347 | 15.4 | 485 | 11.9 | 862 | 18.4 |
| **Truancy** |  |  |  |  |  |  |
| No | 6307 | 84.9 | 2834 | 82.6 | 3473 | 86.9 |
| Yes | 1191 | 15.1 | 634 | 17.4 | 557 | 13.1 |

### Violence and non-fatal injury prevalence

The prevalence of violence and non-fatal injury among Vietnamese adolecents in 2019 was 14.5% (95% CI: 12.6-16.6) and 21.4% (95% CI: 19.8-23.2), respectively. This prevalence was significantly higher in males than in females, with 21.7% (95% CI: 18.1-25.7) vs. 8.3% (95% CI: 7.4-9.5) experiencing violence, and 27.7% (95% CI: 24.6-31.1) vs. 16.1% (95% CI: 14.5-17.8), sustaining non-fatal injuries. The prevalence of violence decreased by age; ranging from 32.4% (95% CI: 24.7-40.2) to 12.6% (95% CI: 8.4-16.8) among males, and 11.1% (95% CI: 7.3-14.9) to 4.6% (95% CI: 3.2-6.0) among females, while differences in non-fatal injury prevalence were not significant between age groups (**Figure 1**).

**Figure 1:**
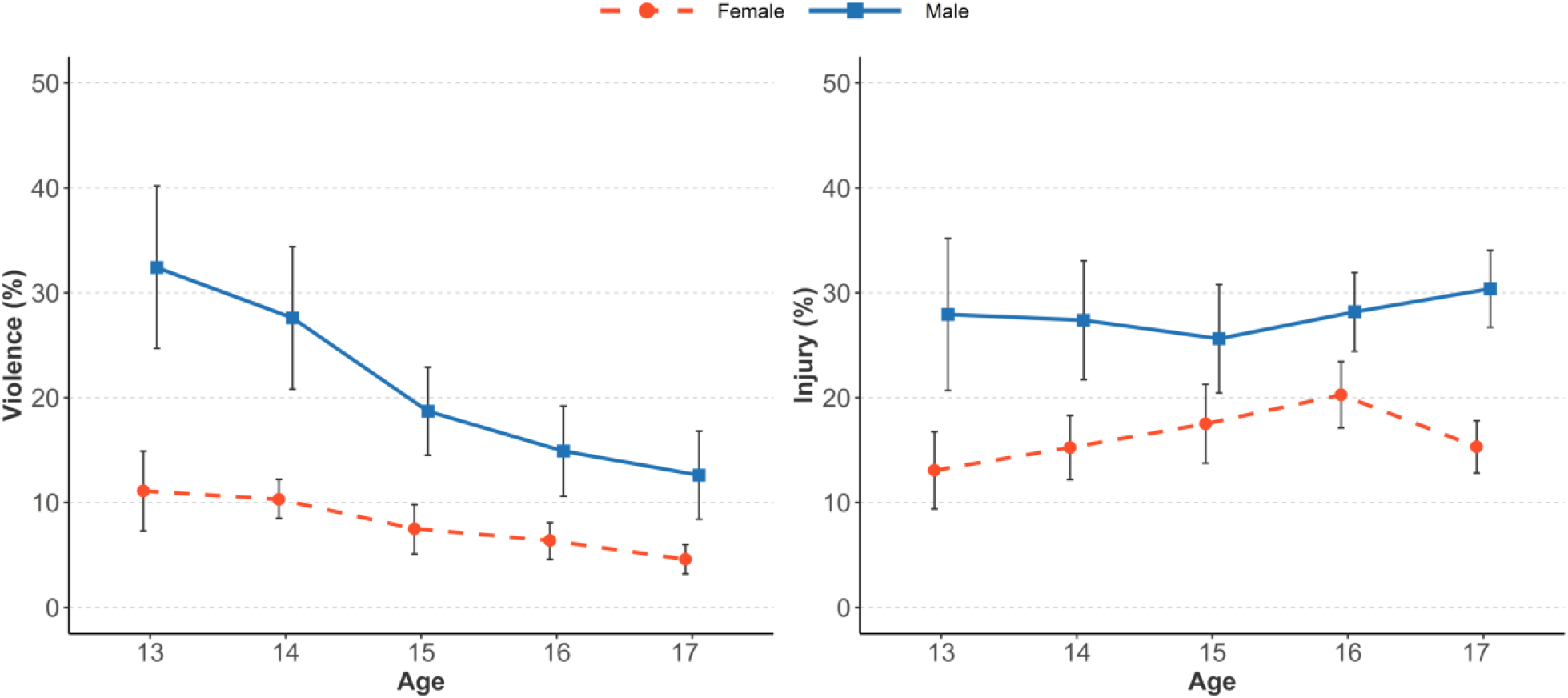
Prevalence of violence and non-fatal injury among in-school adolescent in Vietnam, by age and gender, 2019.

Details of the prevalence of violence and non-fatal injury by student characteristics are available in **Supplemental Table 1**.

### Factors related to violence

**Figure 2** shows the results of multivariable logistic model of factors related to violence among adolescents aged 13-17 in Vietnam in 2019. Conditional on all other factors, drinking alcohol is associated with significantly higher odds of violence among male students (OR = 2.11, 95% CI: 1.45-3.07). Female students living in urban areas or living with neither father nor mother were likely to have higher odds of violence (OR = 1.35, 95% CI: 1.01-1.82, and OR = 1.52, 95% CI: 1.01-2.28, respectively), whereas those with high parental respect had lower odds of violence (OR = 0.58, 95% CI: 0.41-0.82). Smoking cigarettes, loneliness, and suicide attempts were significantly associated with increased odds of violence, while higher age was a protective factor against violence in both genders.

**Figure 2:**
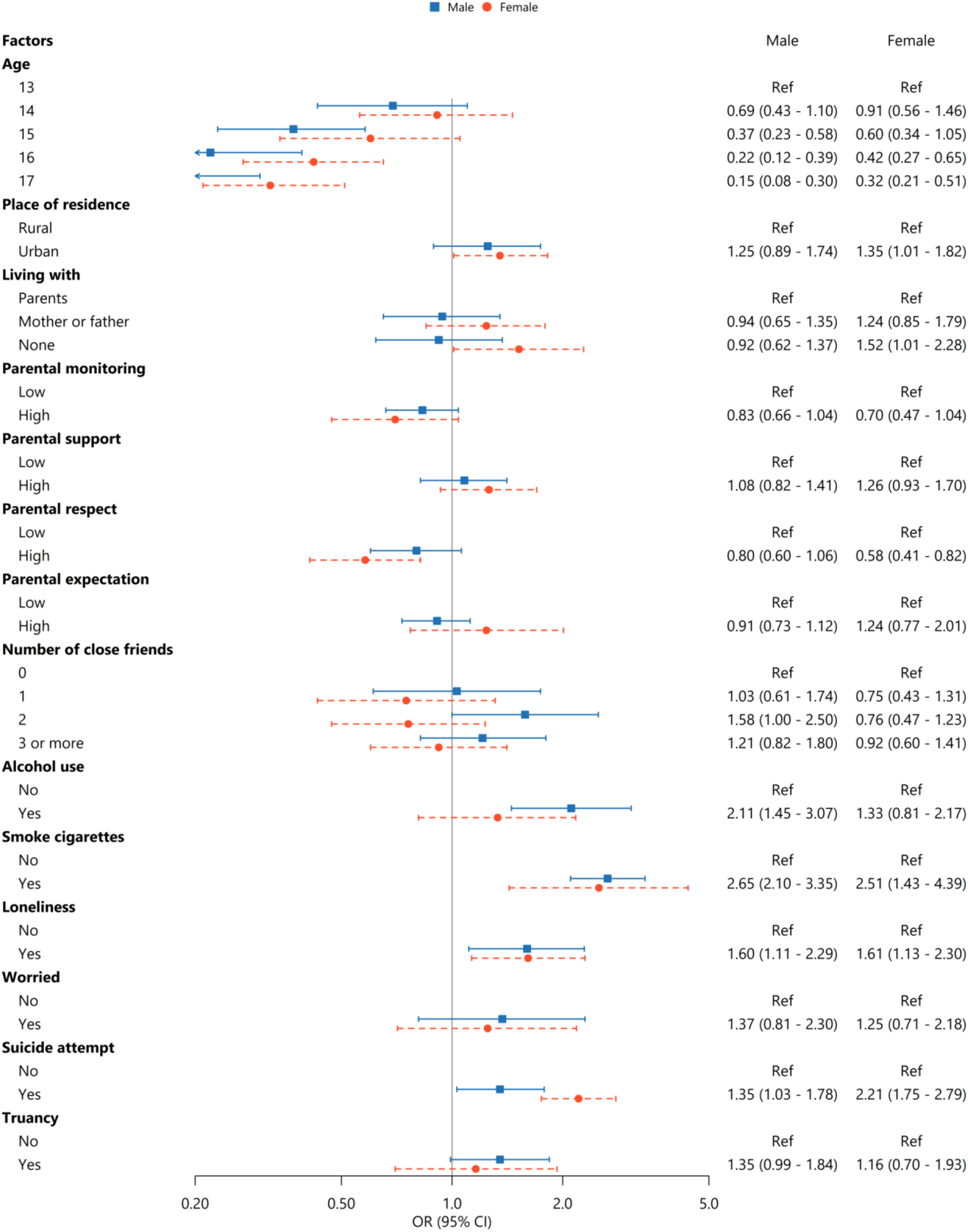
Multivariable logistic model of factors related to violence among in-school adolescent in Vietnam, 2019.

### Factors related to non-fatal injuries

**Figure 3** shows the findings of multivariable logistic model of factors associated with non-fatal injuries among Vietnamese adolescents aged 13-17 in 2019. We found that male students living with neither mother nor father and smoking cigarettes had a higher likelihood of non-fatal injury (OR = 1.30, 95% CI: 1.07-1.58, and OR = 1.43, 95% CI: 1.08-1.88, respectively). Among female students, higher odds of non-fatal injury were found in those who felt worry (OR = 2.06, 95% CI: 1.34-3.17), had attempted suicide (OR = 1.40, 95% CI: 1.13-1.72), and drank alcohol (OR = 1.40, 95% CI: 1.09-1.80). In contrast, female students with parental respect had 40% lower odds of non-fatal injuries (OR = 0.60, 95% CI: 0.42-0.84). In both genders, exposure to violence was a significant risk factor for non-fatal injuries.

**Figure 3:**
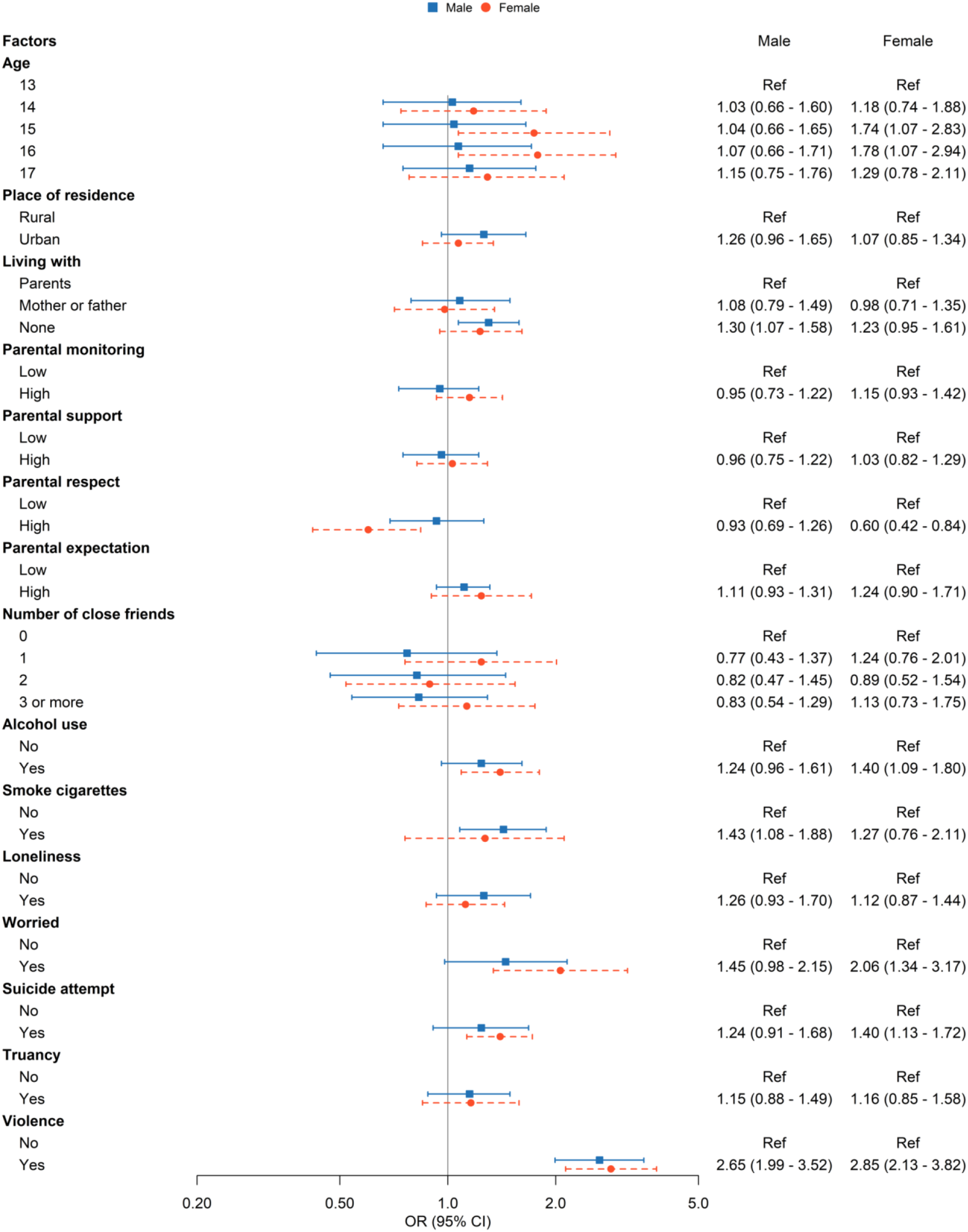
Multivariable logistic model of factors related to non-fatal injury among in-school adolescent in Vietnam, 2019.

## Discussion

In this study, we found that the prevalence of violence among Vietnamese students aged 13-17 is 14.5% and the prevalence of non-fatal injury is 21.4%. Violence and non-fatal injury are both more prevalent among male students compared to female students. Older students are less likely to be involved in violence, although a higher age is not a protective factor for non-fatal injury. Risk factors for violence among students include alcohol use, smoking cigarettes, mental health problems, living in urban areas, and living with neither parent. For non-fatal injury, risk factors include smoking cigarettes, alcohol use, living with neither parent, violence, and mental health problems. It is notable that parental factors have a significant impact on female students regarding both violence and non-fatal injury. Parental respect significantly reduces the odds of violence and non-fatal injury among female students, while living with neither parent increases the risk of violence. Our findings serve as evidence for developing effective prevention and action strategies to mitigate the burden of violence and injury among adolescents in Vietnam and other low- and lower-middle-income countries.

Previous studies have reported that the prevalence of violence is different across countries. Han et al.^3^ found the prevalence of physical attack and physical fighting among 68 low-middle-income countries (LMICs) from 2009 to 2015 to be 35.6 (95% CI: 30.7–40.5) and 36.4 (95% CI: 29.9– 42.9), respectively. The prevalence was lower in high-income countries. For example, the Youth Risk Behavior Survey in 2015 showed that 23.6% (95% CI: 21.6–25.6) of adolescents in the USA had been involved in a physical fight.^20^ The prevalence of physical fighting (defined as at least four incidents of fighting in the previous 12 months) among adolescents in the USA and Northern European countries is less than 6.5%.^21^ The overall prevalence of violence in our study was lower than the findings from other countries. The differences in these prevalence estimates might be explained by the cultural differences between countries in terms of violence. The year when the surveys were conducted might also account for the variation, given that the prevalence of violence in Vietnam in 2013 was 27.2% (95% CI: 24.1–30.5),^22^ which was similar to other countries at that time. A systematic analysis of the global updated data is unfortunately not available; however, it can be observed that the prevalence of violence among Vietnamese students in 2019 reduced significantly compared to 2013. This reduction might be due to Vietnam’s rapid economic growth, and more attention and investment being given to education. A previous study demonstrated the relationship between spending on education and the prevalence of physical fighting among adolescents.^21^ Furthermore, reduction in school violence prevalence in Vietnam would not have been possible without the crucial role of the Ministry of Education and the coordinated effort of school governers and staffs. In 2017, the Ministry of Education issued Decision 5886 which consisted of a comprehensive action plan for prevention and control of school violence.^23^ Indeed, it is very important to integrate violence prevention content not only in formal curriculum but also in extracurricular activities. The decision also emphasizes the importance of communication activies, safe school environment and training for school staffs. It is also notable that the prevalence of violence in our study includes not only students who were victims of violence but also those who were actively involved in physical fights. From our study, it was found that 10.42% (95% CI: 8.98-11.87) of students were physically attacked. Male students were more likely to be victims of violence (15.4%, 95% CI: 12.55-18.26) compared to female students (6.05%, 95% CI:5.28-6.83).

Our results regarding the prevalence of non-fatal injuries among young adolescents are relatively lower than studies from other LMICs. For instance, the prevalence of injury across 68 countries reported by Han et al.^3^ was 42.9% (95% CI: 39.0–46.9), in which the highest prevalence was found in the African region, with 48.1% (95% CI: 40.2–55.9). Street et al.^24^ analyzed the data from 95,811 students aged 13–15 years in 47 LMICs between 2003 and 2012 and found that 39.9% (95% CI: 35.1–44.8) of the participants reported at least one serious injury during the preceding 12 months. Similar to Han et al.,^3^ Street et al. also showed that the highest percentages were in Samoa (72.1%) and six countries in Sub-Saharan Africa.^24^ The differences in prevalence estimates in our study and the above studies might be explained by the differences in the location where the surveys were conducted; the two previous studies included many high-risk regions.^3,24^

Non-fatal injuries among school-aged adolescents in Vietnam in 2019 were also less prevalent compared to 2013 (29.3%).^22^ This result shows great progress in injury prevention in Vietnam. Multisectorial effort among the Ministry of Health, the Ministry of Education, and the Ministry of Labour – Invalids and Social Affairs has led to implementation of many injury prevention activities in the community, focusing on communication to raise awareness, education for school staff and parents, and building safe communities for children and adolescents. However, this prevalence figure was still higher than that reported among adolescents in developed countries; for example, from 9.8% to 7.8% between 2003 and 2011 in Spain.^25^ This finding indicates that despite efforts on injury prevention, serious injury should be considered as a public health priority, particularly in LMICs with limited resources for healthcare.

Regarding gender, our results indicate that male students suffer more violence and injury than female students. These results are consistent with previous studies^17,19,25^ and may be because boys are more physically active than girls, and tend to get involved more easily in competitive activities such as physical fights and games.^26,27^ We observed many common risk factors for violence and non-fatal injury, including the smoking of cigarettes, alcohol use, mental health problems, and living with neither parent, with violence also being a risk factor for injury. These findings are consistent with research from many other countries.^7-9,28-30^ Notably, mental health is a risk factor for non-fatal injuries among female students and for violence in both genders, which accords with previous studies.^29,31^ With the popularity of social platforms and rapid access to a wide range of information, Vietnamese students may face an increased risk of mental health problems, which is particularly worrying given that mental health is also associated with violence and non-fatal injury. Moreover, parents were found to play a crucial role in preventing violence and non-fatal injury among Vietnamese students; this may be explained by Vietnamese family culture, where parents and children have close bonds, and parents are often involved in children’s education, personal development, and even daily activities. Studies from other countries have also indicated that the better and closer the parent-children relationship is, the less likely violence and injury among children will occur.^32-34^ Wright and Fitzpatrick reported that certain aspects of the parent-adolescent relationship, such as trust, involvement, and connection, are protective factors of adolescent violence.^34^ The relationship between parent and child may also impact adolescents in other ways, such as preventing depression and self-harm behaviors, and in turn, preventing violence and injury.^35^ Together, our findings highlight the need for the implementation of effective public health interventions that take into account a comprehensive list of factors to mitigate the burden of violence and injuries among adolescents in Vietnam, including behavioral risk factors, mental health, and adolescent-parent bonding.

### Strengths and limitations

A strength of our study is the large and nationally representative sample, using a standardized study design and research method. The study also used the most recent dataset (GSHS 2019) to provide a timely assessment of the status of violence and non-fatal injury among adolescents in Vietnam, and associated factors. Furthermore, the results from this study account for a wide age range of adolescents (13-17 years old), and are comparable across the country. However, we also recognize some limitations of our study. First, due to its self-reporting nature, the information obtained through the survey may be subject to recall bias. Second, although most adolescents in Vietnam are in school, this study focused only on in-school adolescents, which may not be representative of all adolescents, including those who are not in school. Thirdly, given the cross-sectional design, we could not confirm causal inferences and were not able to evaluate the long-term consequences of violence and injury.

## Conclusion

Despite the reduction in rates of violence and non-fatal injury, the prevalence of these issues is still relatively high among students in Vietnam. Common associated factors of violence and non-fatal injuries include behavioral risk factors, mental health problems, and parental factors. Future policies should consider both individual factors as well as adolescent-parent bonding in order to reduce the burden of violence and injury among adolescents in Vietnam.

## Data Availability

The datasets, codebook used in the current study are not publicly available but are available upon reasonable request

## Declarations

### Ethical approval

All procedures performed in this study were in accordance with the ethical standards of the Institution Review Board of Hanoi University of Public Health (IRB No. 421/2019/YTCC-HD3, dated: 06/08/2019). Written informed consents were obtained from all participants’ parents/guardians before the study. All students participating in this survey were explained about the study. Survey procedures were designed to protect student privacy by allowing for anonymous and voluntary participation. The students had the right to withdraw from the study or refuse to answer any specific questions in the questionnaire without any consequences. All personal information of students was kept confidential, and all information was encrypted.

### Consent for publication

Not applicable

### Authors’ contributions

Conceptualization: LPA; Methodology: LPA, HVM, and KQL; Formal analysis and investigation: LPA and KQL; Writing - original draft preparation: LPA and KQL; Writing - review and editing: HVM, TTTH, MT, NTL, PTQN, LVT, TQB, and KP; Supervision: HVM and TTTH. All authors read and approved the final manuscript.

## References

1. World Health Organization. Youth violence. 2020; https://www.who.int/news-room/fact-sheets/detail/youth-violence. (12 Aug 2020, date last accessed).

2. Akiba M, LeTendre GK, Baker DP, Goesling B. Student Victimization: National and School System Effects on School Violence in 37 Nations. American Educational Research Journal. 2002;39(4):29–53.

3. Han L, You D, Gao X, et al. Unintentional injuries and violence among adolescents aged 12-15 years in 68 low-income and middle-income countries: a secondary analysis of data from the Global School-Based Student Health Survey. The Lancet Child & adolescent health. 2019;3(9):616–26.

4. World Health Organization. Violence and Injury Prevention. 2014; https://www.who.int/violence_injury_prevention/media/news/2015/Injury_violence_facts_2014/en/. (12 Aug 2020, date last accessed).

5. World Health Organization. Maternal, newborn, child and adolescent health. https://www.who.int/maternal_child_adolescent/epidemiology/adolescence/en/. (12 Aug 2020, date last accessed).

6. Molcho M, Harel Y, Pickett W, et al. The epidemiology of non-fatal injuries among 11-, 13- and 15-year old youth in 11 countries: findings from the 1998 WHO-HBSC cross national survey. International Journal of Injury Control and Safety Promotion. 2006;13(4):205–11.

7. Peltzer K, Pengpid S. Injury and social correlates among in-school adolescents in four Southeast Asian countries. International journal of environmental research and public health. 2012;9(8):2851–62.

8. Ohene SA, Johnson K, Atunah-Jay S, Owusu A, Borowsky IW. Sexual and physical violence victimization among senior high school students in Ghana: Risk and protective factors. Social science & medicine (1982). 2015;146:266–75.

9. Senanayake SJ, Gunawardena S, Wickramasinghe S, et al. Prevalence and Correlates of Interpersonal Violence Among In-School Adolescents in Sri Lanka: Results From the 2016 Sri Lankan Global School-Based Health Survey. Asia-Pacific journal of public health. 2019;31(2):147–56.

10. Beck NI, Arif I, Paumier MF, Jacobsen KH. Adolescent injuries in Argentina, Bolivia, Chile, and Uruguay: Results from the 2012-2013 Global School-based Student Health Survey (GSHS). Injury. 2016;47(12):2642–49.

11. Denny VC, Cassese JS, Jacobsen KH. Nonfatal injury incidence and risk factors among middle school students from four Polynesian countries: The Cook Islands, Niue, Samoa, and Tonga. Injury. 2016;47(5):1135–42.

12. Peltzer K. Injury and social determinants among in-school adolescents in six African countries. Injury prevention: journal of the International Society for Child and Adolescent Injury Prevention. 2008;14(6):381–8.

13. Peltzer K, Pengpid S. Unintentional Injuries and Psychosocial Correlates among in-School Adolescents in Malaysia. International journal of environmental research and public health. 2015;12(11):14936–47.

14. Muula AS, Siziya S, Rudatsikira E. Prevalence and socio-demographic correlates for serious injury among adolescents participating in the Djibouti 2007 Global School-based Health Survey. BMC research notes. 2011;4:372.

15. Peyton RP, Ranasinghe S, Jacobsen KH. Injuries, Violence, and Bullying Among Middle School Students in Oman. Oman medical journal. 2017;32(2):98–105.

16. Pierobon M, Barak M, Hazrati S, Jacobsen KH. Alcohol consumption and violence among Argentine adolescents. Jornal de pediatria. 2013;89(1):100–7.

17. Pengpid S, Peltzer K. Unintentional injuries and socio-psychological correlates among school-going adolescents in four ASEAN countries. International journal of general medicine. 2019;12:263–71.

18. Vietnam General Statistics Officice. Average population by location. 2020; https://www.gso.gov.vn/default.aspx?tabid=722. (12 Aug 2020, date last accessed).

19. Peltzer K, Pengpid S. Nonfatal Injuries and Psychosocial Correlates among Middle School Students in Cambodia and Vietnam. International journal of environmental research and public health. 2017;14(3).

20. Centers for Disease Control and Prevention. Youth Online: High school YRBS. https://nccd.cdc.gov/youthonline/app/. (12 Aug 2020, date last accessed).

21. Elgar FJ, McKinnon B, Walsh SD, et al. Structural Determinants of Youth Bullying and Fighting in 79 Countries. The Journal of adolescent health: official publication of the Society for Adolescent Medicine. 2015;57(6):643–50.

22. World Health Organization. Global school-based student health survey Vietnam: Country fact sheet. World Health Organization; 2013.

23. Ministry of Education of Vietnam. Decision 5886: Programs on preveting school violence in primary, high school, and continuing education for the period 2017-2021. Hanoi, Vietnam2017.

24. Street EJ, Jacobsen KH. Injury incidence among middle school students aged 13-15 years in 47 low-income and middle-income countries. Injury prevention: journal of the International Society for Child and Adolescent Injury Prevention. 2016;22(6):432–6.

25. Alonso-Fernández N, Jiménez-Garcí a R, Alonso-Fernández L, Hernández-Barrera V, Palacios-Ceña D. Unintentional injuries and associated factors among children and adolescents. An analysis of the Spanish National Health Survey. International Journal of Public Health. 2017;62(9):961–9.

26. Patel D, Magnusen E, Sandell JM. Prevention of unintentional injury in children. Paediatrics and Child Health. 2017;27(9):420–6.

27. Zoni AC, Domínguez-Berjón MF, Esteban-Vasallo MD, Regidor E. Injuries treated in primary care in the Community of Madrid: analyses of electronic medical records. Gaceta sanitaria. 2014;28(1):55–60.

28. Bala MO, Chehab MA, Al-Dahshan A, Saadeh S, Al Khenji A. Violence among Adolescents in Qatar: Results from the Global School-based Student Health Survey, 2011. Cureus. 2018;10(7):e2913.

29. Kelishadi R, Babaki AE, Qorbani M, et al. Joint Association of Active and Passive Smoking with Psychiatric Distress and Violence Behaviors in a Representative Sample of Iranian Children and Adolescents: the CASPIAN-IV Study. International journal of behavioral medicine. 2015;22(5):652–61.

30. Muula AS, Siziya S, Rudatsikira E. Self-inflicted serious injuries among adolescents in Zambia. Tanzania journal of health research. 2013;15(1):51–57.

31. Pengpid S, Peltzer K. High prevalence of unintentional injuries and socio-psychological correlates among school-going adolescents in Timor-Leste. International journal of adolescent medicine and health. 2020.

32. Howard KS, Lefever JE, Borkowski JG, Whitman TL. Fathers’ influence in the lives of children with adolescent mothers. Journal of family psychology: JFP: journal of the Division of Family Psychology of the American Psychological Association (Division 43). 2006;20(3):468–76.

33. Musitu Ochoa G, Estévez Lopez E, Emler NP. Adjustment problems in the family and school contexts, attitude towards authority, and violent behavior at school in adolescence. Adolescence. 2007;42(168):779–94.

34. Wright DR, Fitzpatrick KM. Social Capital and Adolescent Violent Behavior: Correlates of Fighting and Weapon Use among Secondary School Students. Social Forces. 2006;84(3):1435–53.

35. Silver ME, Field TM, Sanders CE, Diego M. Angry adolescents who worry about becoming violent. Adolescence. 2000;35(140):663–9.

